# Evaluation of safety and efficacy of autologous oral mucosa-derived epithelial cell sheet transplantation for prevention of anastomotic restenosis in congenital esophageal atresia and congenital esophageal stenosis: three case studies

**DOI:** 10.1101/2022.09.04.22279376

**Authors:** Akihiro Fujino, Yasushi Fuchimoto, Teizaburo Mori, Motohiro Kano, Yohei Yamada, Michinobu Ohno, Yoshiyuki Baba, Nobutaka Isogawa, Katsuhiro Arai, Takako Yoshioka, Makoto Abe, Nobuo Kanai, Ryo Takagi, Masanori Maeda, Akihiro Umezawa

**Author notes:** Correspondence should be directed to: Yasushi Fuchimoto M.D., Ph.D., Department of Pediatric Surgery, International University of Health and Welfare School of Medicine, 852, Hatakeda, Narita, Chiba, 286-8686, Japan.

## Abstract

**Background:** We performed the first autologous oral mucosa-derived epithelial cell sheet transplantation therapy in a patient with refractory postoperative anastomotic stricture in congenital esophageal atresia (CEA) and confirmed its safety. In this study, patients with CEA and congenital esophageal stenosis (CES) were newly added as subjects to further evaluate the safety and efficacy of cell sheet transplantation therapy.

**Methods:** Epithelial cell sheets were prepared from the oral mucosa of the subjects and transplanted into esophageal tears created by endoscopic balloon dilatation (EBD). The safety of the cell sheets was confirmed by quality control testing, and the safety of the transplantation treatment was confirmed by 48-week follow-up examinations.

**Results:** Subject 1 had a stenosis resected because the frequency of EBD did not decrease after the second transplantation. Histopathological examination of the resected stenosis revealed marked thickening of the submucosal layer. Subject 2 did not require EBD for more than 18 months after transplantation, and Subject 3 did not require EBD for at least 9 months after transplantation, during which time they were able to maintain a normal diet by mouth.

**Conclusions:** Subject 2 was free of EBD for a long period of time after transplantation, confirming that cell sheet transplantation therapy is clearly effective in some cases. In the future, it is necessary to study more cases; develop new technologies such as an objective index to evaluate the efficacy of cell sheet transplantation therapy and a device to achieve more accurate transplantation; identify cases in which the current therapy is effective; find the optimal timing of transplantation; and clarify the mechanism by which the current therapy improves stenosis.

**Trial registration:** UMIN, UMIN000034566, registered 19 October 2018, https://upload.umin.ac.jp/cgi-open-bin/ctr_e/ctr_view.cgi?recptno=R000039393.

## Background

Postoperative anastomotic stenosis in congenital esophageal atresia (CEA) and congenital esophageal stenosis (CES) has been reported to occur in 30% to 50% of cases [1–3]. The main symptom of anastomotic stenosis is dysphagia, resulting in feeding difficulties such as choking on food or difficulty in swallowing saliva. Some of these patients are treated with endoscopic balloon dilatation (EBD) with or without local steroid injections and gradually improve. However, there are many cases of refractory anastomotic stenosis that require repeated EBD. These patients not only suffer physical effects such as malnutrition and poor growth, but also often lose the opportunity to enjoy meals with others. In addition, EBD requires hospitalization, which is time-consuming, physically restricting, and emotionally distressing. For patients who require ongoing treatment for restenosis, resection and re-anastomosis of the stenosis or placement of an absorbable stent are recommended [1]. Re-anastomosis is chosen in severely intractable cases. However, in those cases, the upper and lower esophagus had been stitched together and exhibit severe stretching in the initial anastomosis, resulting in suture failure, and fibrotic scar stenosis, or anastomosis that requires a substitute esophagus. Re-anastomosis itself could be a life-threatening risk. Furthermore, re-stenosis may occur even after re-anastomosis, so a safer, and minimally invasive treatment is desirable. Even if restenosis occurs to some extent, but the lumen of the stenosis is kept at a certain size and/or the stenosis is flexible, there is a possibility that the patient may keep eating and drinking orally. From this point of view, treatment to at least alleviate restenosis is desired.

To improve this situation for these patients, a new regenerative therapy using somatic stem cells was devised, in which autologous oral mucosa-derived epithelial cell sheets prepared from the patient’s oral mucosa were transplanted into a lacerated wound after EBD surgery [4]. This is based on “cell sheet engineering” proposed by Okano et al. at the Institute of Advanced Biomedical Engineering and Science (ABMES), Tokyo Women’s Medical University. Endoscopic submucosal dissection (ESD) is performed for superficial esophageal squamous cell carcinoma (ESCC) in adults, but extensive dissection of the esophageal mucosa and submucosa can cause esophageal stenosis after surgery. Okano et al. reported that transplantation of autologous oral mucosa-derived epithelial cell sheets into ulcerated areas after ESD effectively prevented esophageal stenosis [5–7]. Based on these previous studies, we first confirmed the safety and efficacy of cell sheet transplantation in a preclinical study using a porcine model in order to apply cell sheet transplantation to refractory anastomotic stenosis after surgery for congenital esophageal atresia [8]. Then, we conducted the first clinical trial in humans [4]. Autologous oral mucosa-derived cell sheets were produced from the subject’s oral epithelial tissue, and the sheets were transplanted into a lacerated wound after EBD surgery using a newly developed pediatric transplantation device. The safety of this treatment in humans was confirmed by quality control testing of the cell sheets and follow-up examinations for 48 weeks after transplantation. In this study, the safety and efficacy of cell sheet transplantation therapy for refractory esophageal anastomotic stenosis were evaluated in additional subjects.

## Methods

### Subject information

This study included patients between 1 and 30 years of age with postoperative anastomotic restenosis of CEA and CES who had repeated restenosis after at least 5 balloon dilatations. Three subjects have been treated so far (Table 1).

**Table 1.** Information on subjects

|  | Subject 1 | Subject 2 | Subject 3 |
| --- | --- | --- | --- |
| Age at transplant | Late teens | Late teens | Early teens |
| Sex | Male | Male | Male |
| Height (cm) just before transplantation | 165.3 | 154.7 | 154.9 |
| Weight (kg) just before transplantation | 43.7 | 38.2 | 43.0 |
| Diagnosis | Esophageal atresia (Type B) | Esophageal atresia (Type A) | Esophageal stenosis |
| EBD frequency before transplant | Every 2 or 3 weeks | Every 3 months | 2 or 3 times a year |

### Fabrication of oral mucosal epithelial cell sheets

In accordance with our previous study, cultured autologous oral mucosal epithelial cell sheets were fabricated by using buccal mucosal tissue and serum derived from the patient at the cell processing facility (CPF) of CellSeed Inc. [4]. In brief, buccal mucosal tissue and blood were harvested from the patients at the National Center for Child Health and Development (NCCHD) and transported to the CPF of CellSeed. Autologous serum of the patient was prepared by centrifugation and used as a supplement of culture medium. The oral mucosal cells were prepared by treatment with 1,000 U/mL Dispase I (Godo Shusei, Chiba, Japan) and 2.5 mg/mL trypsin and 380 µg/mL ethylenediamine tetraacetic acid tetrasodium (EDTA) salt dihydrate (Gibco/Thermo Fisher Scientific, MA, USA). The disaggregated oral mucosal cells were seeded onto temperature-responsive cell culture inserts (CellSeed, Tokyo, Japan) at a density of 8.6 to 12.0 x10^4^ cells/cm^2^ and cultured in keratinocyte culture medium (KCM) at 37°C under 5% CO_2_ in the cell culture incubator. The composition of KCM is shown in a previous study [4]. After the cultivation for 16 days, the epithelial cells were transported to the NCCHD from CPF of CellSeed at 37°C and transplanted on wound of esophageal mucosa right after EBD in the operation room. (Table 2)

**Table 2.**
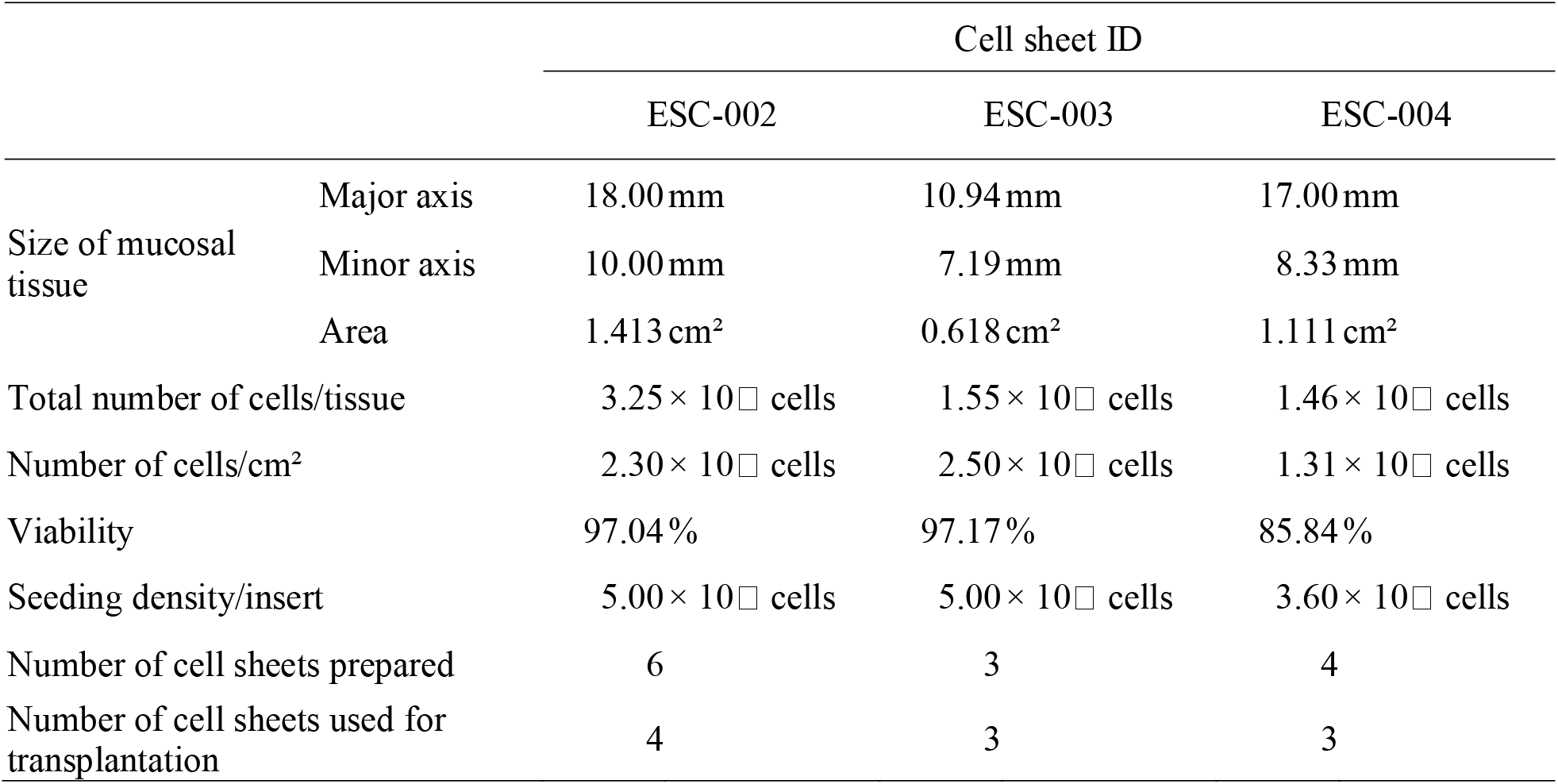
Preparation of oral mucosal epithelial cell sheets

### Quality control tests

In accordance with our previous study, quality control tests were carried out by CellSeed Inc. before transplantation of cultured autologous oral mucosal epithelial cell sheets [4]. Culture media collected during cultivation of the epithelial cells were subjected to sterility tests by (1) cultivation for aerobic and anaerobic bacteria and fungi, (2) endotoxin detection by a limulus amoebocyte lysate assay, and (3) mycoplasma testing by real-time qPCR. Quantification for cellular density, viability, and percentage of epithelial cell in the epithelial cell sheets were also implemented using protocols similar to a previous study [4].

### Endoscopic balloon dilatation (EBD) for anastomotic stenosis

EBD was performed on each subject as in the first transplantation on subject 1 [4]. An esophageal balloon dilation catheter (CRE Fixed Wire, Boston Scientific Corporation, Natick, Massachusetts, USA) were inserted under endoscopic (XQ240, Olympus, Tokyo, Japan) observation. The balloon was inflated by radiopaque contrast medium injection. In Subject 1, the balloon size was increased to 18 mm, 19 mm, and 20 mm, and kept dilated for 180 seconds each time. Subject 2 used a 13.5 mm balloon for dilation for 180 seconds 3 times. Subject 3 used a 20 mm balloon and 180 second dilation was performed three times.

### Endoscopic cell sheet transplantation using a dedicated transplant device

Cell sheets were transplanted into each subject under the same conditions as for the first transplantation in subject 1 [4]. Using a pediatric cell sheet transplantation device developed for this study, epithelial cell sheets were applied to the area of mucosal defect created by EBD avoiding overlap each other. Four, 3, and 3 cell sheets (sheet ID: ESC-002, -003, and -004, respectively) were used for subject 1, 2 and 3, respectively (Table 2).

### Follow-up examination

To assess the safety and the efficacy, scheduled follow-up examinations were performed for 48 weeks after transplantation of cell sheets in each subject under the same conditions as in the first transplantation in subject 1 [4]. The follow-up examinations included eight evaluations: (1) measurement of vital signs, (2) determination of dysphagia and choking on food, (3) blood tests, (4) x-rays, (5) endoscopy for stenosis, (6) endoscopy for sheet fixation and epithelialization, (7) esophagography for stricture, and (8) adverse events (Supplemental Table 1). If symptoms such as dysphagia occurred, endoscopy and esophagography were performed as needed.

## Results

### Fabrication of epithelial cell sheets

Autologous epithelial cells were prepared from buccal mucosal tissues of the subjects, and the epithelial cells were seeded onto temperature-responsive culture inserts at a density of 10.8 ± 1.92 × 10^4^ cells/cm^2^ for fabricating epithelial cell sheets. The results of cell preparation are shown in Table 2. During the cultivation of the epithelial cells, culture media after medium replacement were collected and used for sterility and mycoplasma tests. After 15 days of cultivation, one or two cell culture inserts with the cultured epithelial cells was used for quality control tests in each of three cases. The results of these tests are shown in Table 3 and Supplemental Figure 1. In all cases, the epithelial cell sheets met the criteria for use in transplantation.

**Table 3.**
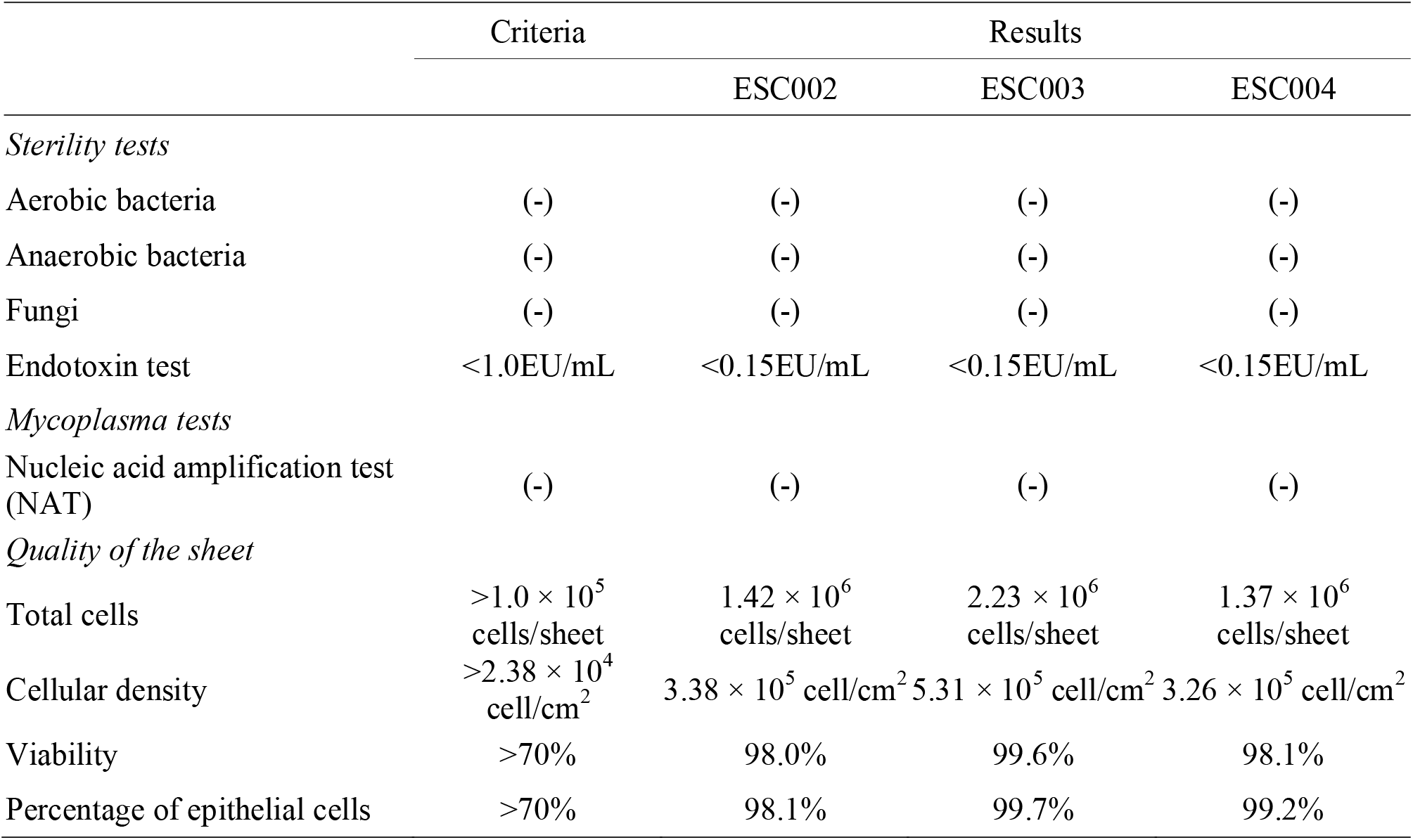
Quality control tests for oral mucosal epithelial cell sheets

|  | Criteria | Results |  |  |
| --- | --- | --- | --- | --- |
|  |  | ESC002 | ESC003 | ESC004 |
| *Sterility tests* |  |  |  |  |
| Aerobic bacteria | (-) | (-) | (-) | (-) |
| Anaerobic bacteria | (-) | (-) | (-) | (-) |
| Fungi | (-) | (-) | (-) | (-) |
| Endotoxin test | <1.0EU/mL | <0.15EU/mL | <0.15EU/mL | <0.15EU/mL |
| *Mycoplasma tests* |  |  |  |  |
| Nucleic acid amplification test (NAT) | (-) | (-) | (-) | (-) |
| *Quality of the sheet* |  |  |  |  |
| Total cells | $>1.0 \times 10^5$ cells/sheet | $1.42 \times 10^6$ cells/sheet | $2.23 \times 10^6$ cells/sheet | $1.37 \times 10^6$ cells/sheet |
| Cellular density | $>2.38 \times 10^4$ cell/cm<sup>2</sup> | $3.38 \times 10^5$ cell/cm<sup>2</sup> | $5.31 \times 10^5$ cell/cm<sup>2</sup> | $3.26 \times 10^5$ cell/cm<sup>2</sup> |
| Viability | >70% | 98.0% | 99.6% | 98.1% |
| Percentage of epithelial cells | >70% | 98.1% | 99.7% | 99.2% |

### Transplantation of epithelial cell sheets and progress in subject 1

A male in his late teens with postoperative anastomotic stenosis of esophageal atresia type B was the first subject, for whom a second epithelial cell sheet transplantation was performed (Table 1, Figure 1). Subject 1 had restenosis after the first cell sheet transplantation and underwent EBDs (Figure 1A). Since EBD was required every 2 to 4 weeks thereafter, a second transplant was performed 13 months after the first transplant. The length of the stenosis was measured on contrast-enhanced images as 20.8 mm to 36.4 mm, depending on the site (Figure 1B, C). The EBD before transplantation resulted in a circumferential laceration, and a longitudinal laceration extending the entire length of the stenosis at a side, and detachment of the esophageal mucosa. (Figure 1D). Four of the cell sheets (ESC-002) were used for transplantation (Table 2). As seen after the first transplantation, the subject experienced improvement in swallowing food and drink without blockage for some time after the second transplantation. However, the frequency of EBD returned quickly to the pre-transplantation level, and surgical resection of the stenosis was performed 4 months after the second transplantation. Histopathological examination of the resected esophageal stenotic site revealed prominent fibrosis and thickening of the submucosa (HE staining) (Figure 1E, F). The thickness of submucosal layer was 1.8 to 2.0 mm. Desmin staining and alpha-smooth muscle actin (α-SMA) staining revealed lack of muscular continuity in the stenotic site. An accumulation of myofibroblasts was observed in the submucosa of the area of smooth muscle defect.

**Figure 1.**
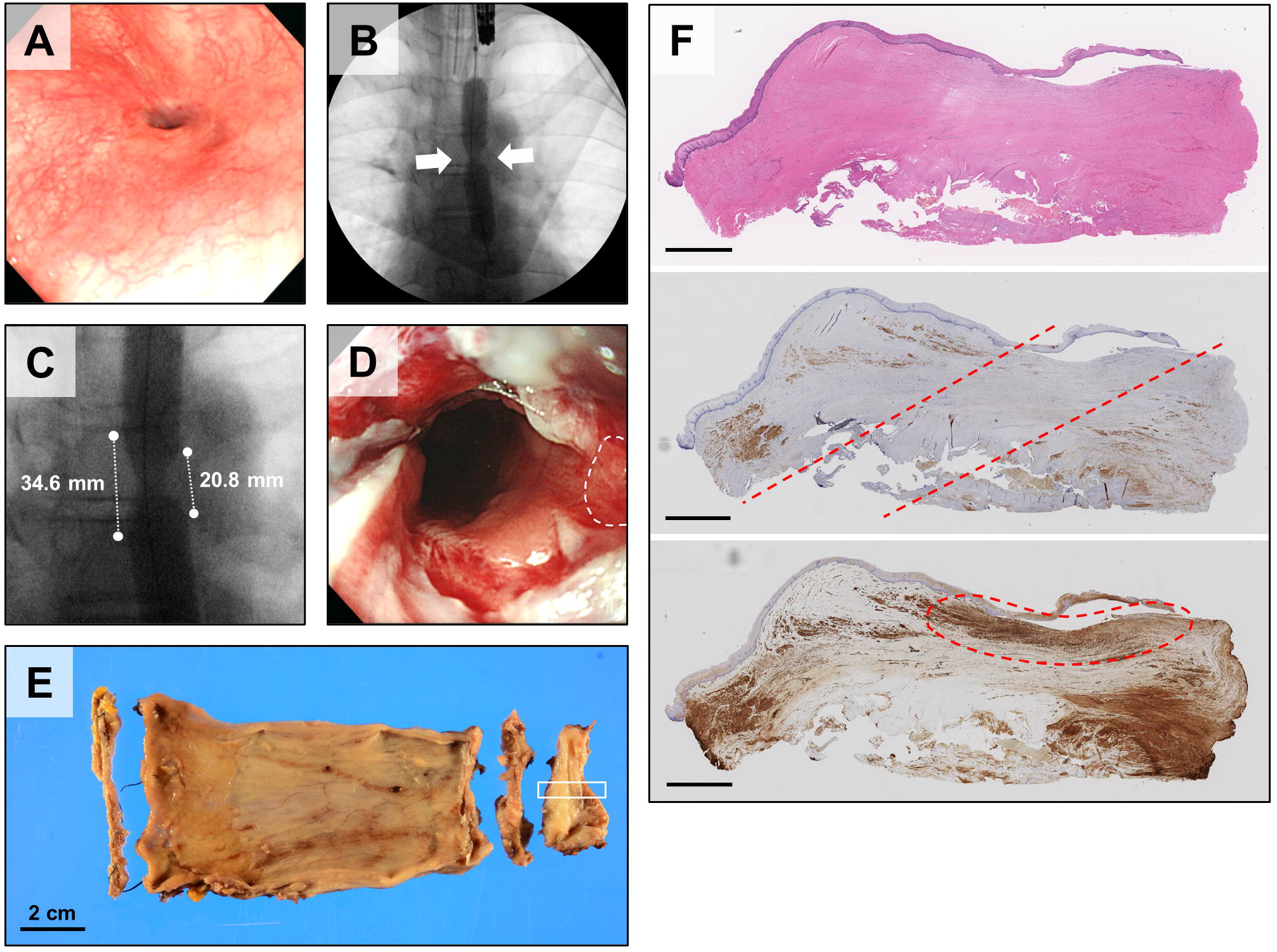
Second epithelial cell sheet transplantation in subject 1. A) Endoscopic image of the transplanted area approximately 3 weeks (23 days) after the first epithelial cell sheet transplantation. The first EBD after transplantation was performed on the same day. B) Contrast esophagography during EBD. Arrows indicate the stenotic area. The balloon was filled with contrast at 1 atm of internal pressure. C) Enlarged image of the esophageal stenosis. The dotted line on the left side is approximately 34.6 mm and the right side is 20.8 mm. D) Endoscopic image immediately after the second cell sheet transplantation. The cell sheet was transplanted in the mucosal defect area indicated by the dotted line. E) Macroscopic view of the resected postoperative anastomotic stenosis of esophageal atresia (right: mouth side, left: stomach side). F) Histology of the stenotic region surrounded by the white line in figure 1E. The submucosal layer was thickened with fibrotic tissue (HE staining, upper panel). In the area between the dotted lines, a lack of continuity of the muscle layer was observed (desmin staining, middle panel). In the area enclosed by the dotted line, an accumulation of myofibroblasts was observed in the submucosa of the smooth muscle defect (α-SMA staining, lower panel). Scale bar is 2 mm.

### Transplantation of epithelial cell sheets and progress in subject 2

The second subject was a male in his late teens with postoperative anastomotic stenosis of esophageal atresia type A (Table 1, Figure 2). The length of the stenosis was measured on contrast-enhanced images as 11.5 mm to 15.1 mm depending on the site (Figure 2B, C). The laceration caused by EBD at transplantation was localized to a single site of laceration over the entire length of the stenosis and a detachment of the esophageal mucosa in the vicinity (Figure 2D). Three cell sheets (ESC-003) were transplanted (Table 2, Figure 2E).

**Figure 2.**
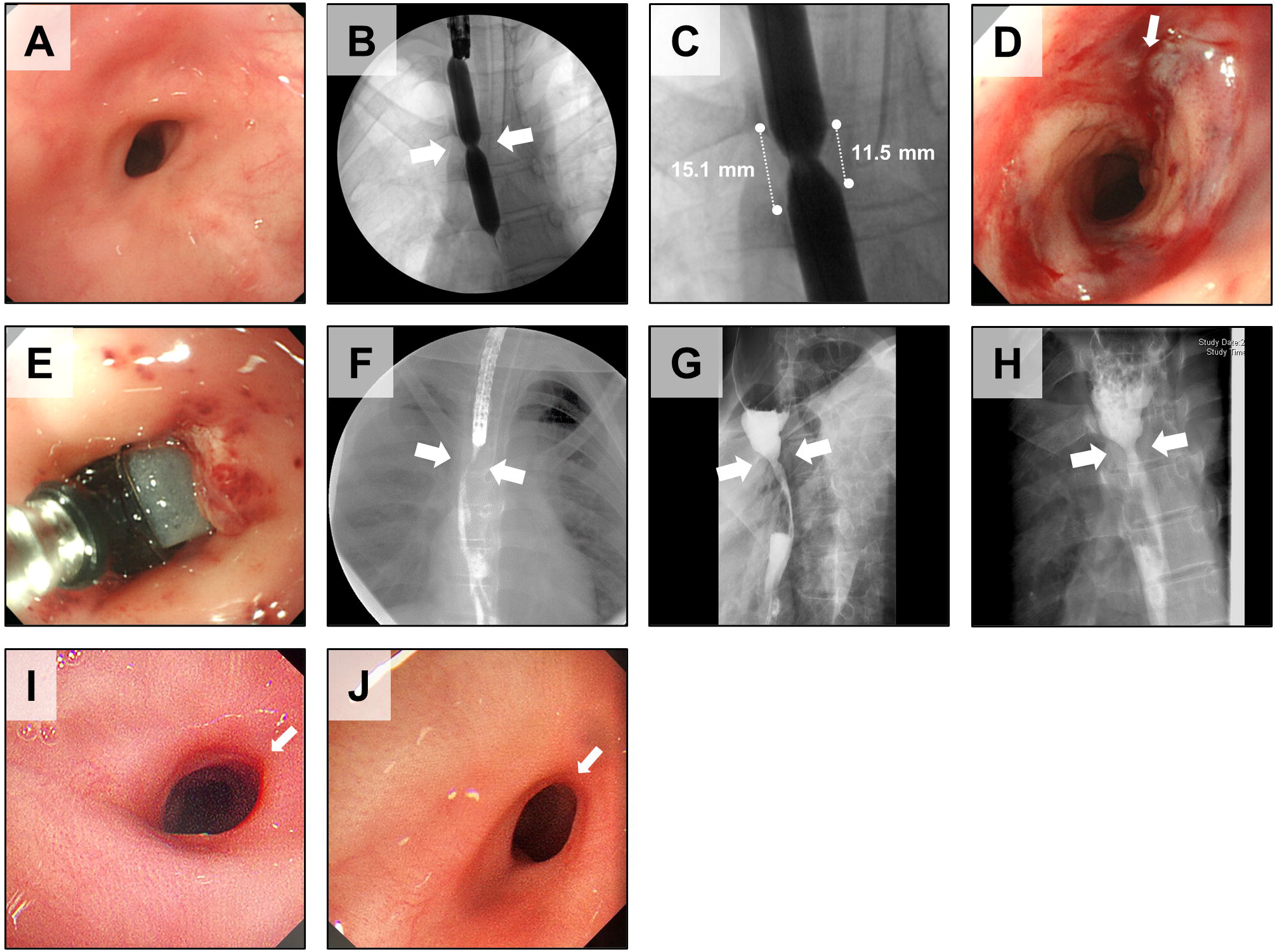
Epithelial cell sheet transplantation into subject 2. A) Endoscopic image of the stenosis just before EBD at cell sheet transplantation. B) Contrast esophagography during EBD just before cell sheet transplantation. Arrows indicate the stenosis. The balloon was filled with contrast at 1 atm of internal pressure. C) Enlarged image of the esophageal stenosis. The dotted line on the left is approximately 15.1 mm and the right is 11.5 mm. D) Endoscopic image after EBD just before cell sheet transplantation. Arrows indicate the location of the laceration caused by EBD. E) The cell sheets were applied to the mucosa dehiscence above the laceration using the transplantation device. F) Contrast esophagography before EBD at cell sheet transplantation. Arrows indicate anastomotic stenosis. G) Contrast esophagography approximately one month (39 days) after cell sheet transplantation. Arrows indicate the stenosis. H) Contrast esophagography approximately 5 months (154 days) after cell sheet transplantation. Arrows indicate the stenosis. I) Endoscopic image of the stenosis approximately 5 months (154 days) after cell sheet transplantation. Arrows indicate the location of the laceration caused by EBD immediately before transplantation. J) Endoscopic image of the stenosis approximately 12 months (348 days) after cell sheet transplantation. Arrows indicate the location of the laceration caused by EBD immediately before transplantation.

Comparison of the diameter at the esophageal stricture in pre- and post-transplantation contrast esophagographies showed no noticeable change (Figure 2F-H). Endoscopic examination at 1, 3, and 6 months after transplantation revealed that the folds behind the stenosis were more easily visible than before transplantation, and the stenosis was very soft (Figures 2I, J). The subject experienced a clear improvement in swallowing food and drink without blockage after transplantation. Before transplantation, the subject was only able to consume a paste diet three months after EBD. However, after transplantation, the subject was able to consume a normal diet for more than one and a half years.

### Transplantation of epithelial cell sheets and progress in subject 3

The third subject was a male in his early teens with postoperative anastomotic stenosis of a congenital esophageal stenosis (Table 1, Figure 3). The length of the stenosis was measured on contrast-enhanced images as 21.8 mm to 22.7 mm depending on the site (Figure 3B, C). The EBD at transplantation caused a circumferential laceration, and a longitudinal laceration extending the entire length of the stenosis on one side, and detachment of the esophageal mucosa. (Figure 3D). Three cell sheets (ESC-004) were transplanted (Table 2, Figure 3E).

**Figure 3.**
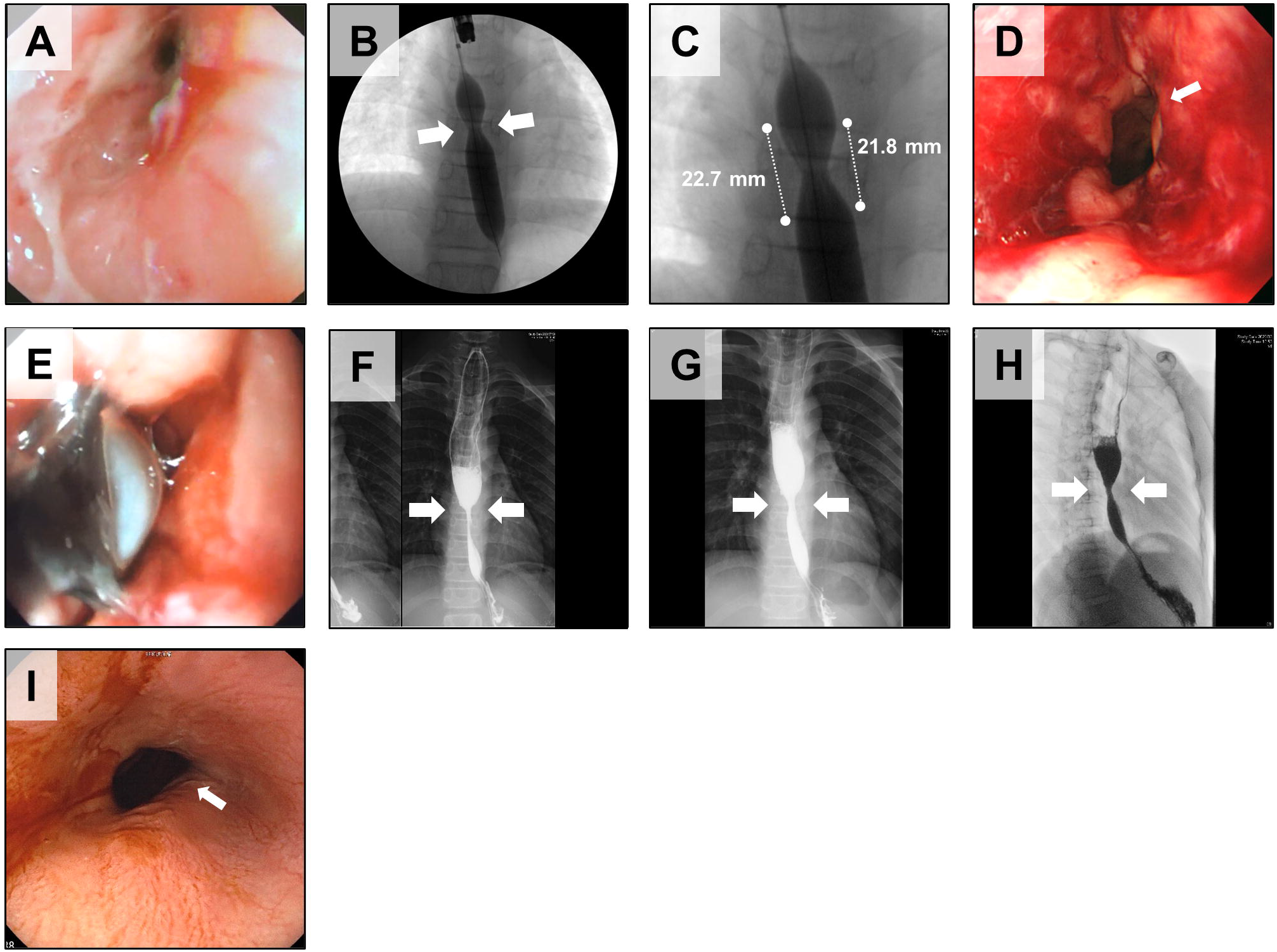
Epithelial cell sheet transplantation into subject 3. A) Endoscopic image of the stenosis just before EBD at cell sheet transplantation. B) Contrast esophagography during balloon dilation just before cell sheet transplantation. C) Arrows indicate the stenosis. The balloon was filled with contrast at 1 atm of internal pressure. D) Enlarged image of the esophageal stenosis. The dotted line on the left is approximately 22.7 mm and the right is 21.8 mm. E) Endoscopic image after balloon dilation just before cell sheet implantation. Arrows indicate the location of the laceration caused by balloon dilation. F) The cell sheets were attached to the mucous dehiscence above the laceration using the transplantation device. G) Contrast esophagography approximately one month before cell sheet transplantation. Arrows indicate anastomotic stenosis. H) Contrast esophagography approximately 3 months (91 days) after cell sheet transplantation. Arrows indicate stenosis. I) Contrast esophagography approximately five and a half months (166 days) after cell sheet transplantation. Arrows indicate the stenosis. J) Endoscopic image of the stenosis approximately one month (28 days) after cell sheet transplantation. Arrows indicate the location of the laceration caused by balloon dilation just before transplantation.

Comparison of the diameter at the esophageal stricture in pre- and post-transplantation contrast esophagographies showed no noticeable change, although the diameter of the stricture appeared to be slightly larger after transplantation (Figure 3F-I). The subject experienced improvement in swallowing food and drink without blockage after transplantation. The time required for a single meal was reduced from 2 hours to about 30 minutes.

### EBDs before and after epithelial cell sheet transplantation

The time course of EBD before and after cell sheet transplantation for each subject is shown in figure 4. In case 1, EBDs were performed less frequently for 6 months after the first transplantation but returned to the same level of frequency as before transplantation; after the second transplantation, the frequency of EBD did not decrease, and the stenosis was finally resected surgically. In case 2, EBDs were performed every 3 months before transplantation, but were not performed more than 18 months after transplantation. In case 3, EBDs were performed 2 to 3 times a year before transplantation but was not performed more than 9 months after transplantation.

**Figure 4.**
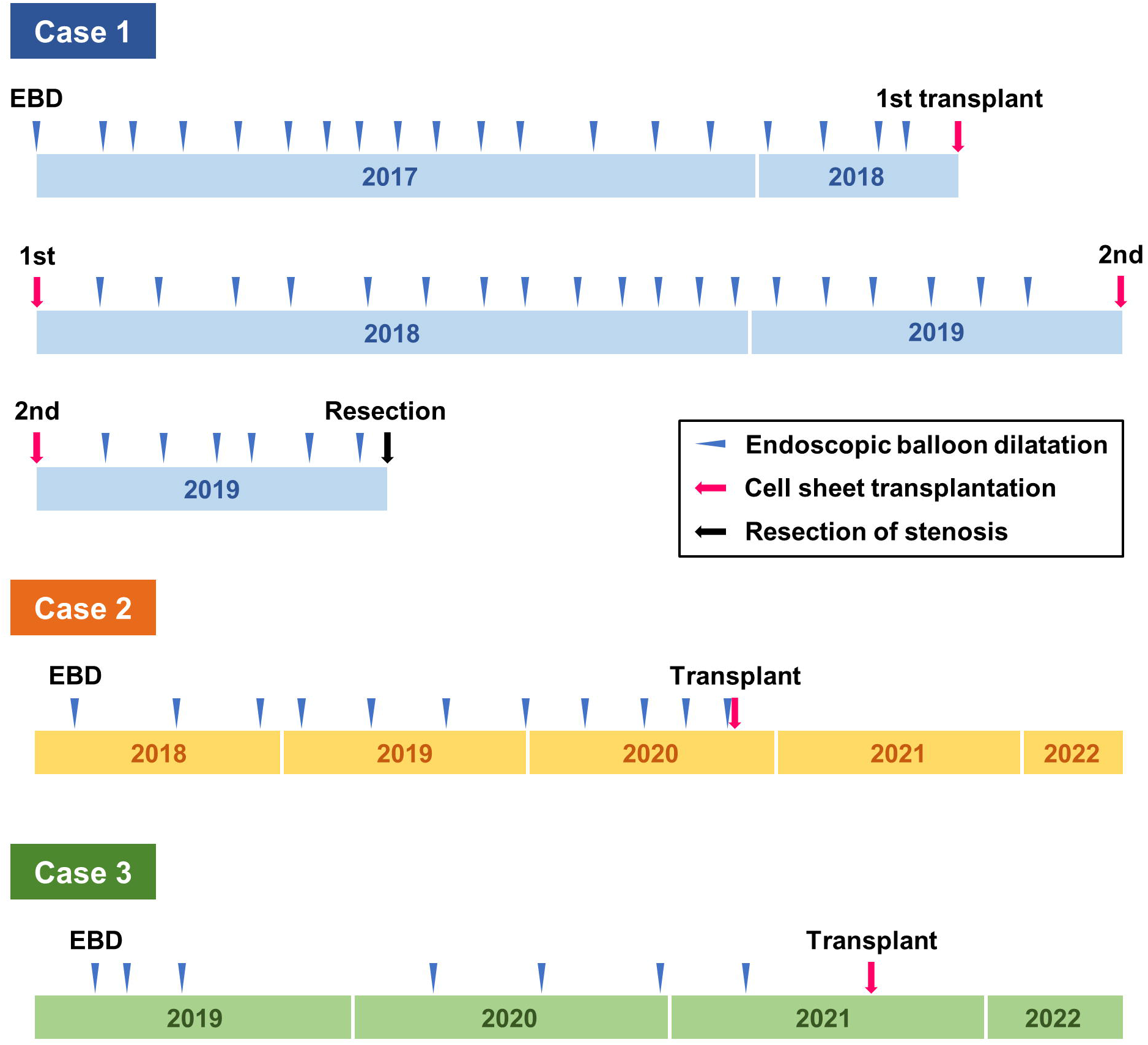
Endoscopic balloon dilatation before and after cell sheet transplantation in each case. The time axis shows the status of EBD before and after cell sheet transplantation in Subjects 1-3. Blue triangles indicate EBD, red arrows indicate cell sheet transplantation, and a black arrow indicates resection of the stenosis. The distance between triangles and arrows indicates the interval between procedures.

### Confirmation of safety

Appropriate follow-up examinations were performed on all subjects. (Subject 3 is ongoing.) All physical and laboratory tests showed no abnormalities, confirming the safety of the transplant.

## Discussion

We reported the first in-human regenerative therapy for postoperative anastomotic stenosis in CEA by transplantation of autologous oral mucosa-derived epithelial cell sheets into the laceration after EBD to prevent restenosis [4]. Here, we reported the progress since then and add new cases.

### Effect of the cell sheet transplantation for the post-anastomotic stenosis of CEA and CES

Subject 1 had undergone EBD every 2 to 3 weeks more than 100 times in total before the first transplantation [4]. This was a particularly severe case of refractory anastomotic stenosis with a long, circumferential stenosis. This subject had a decrease in frequency of EBD for 6 months after the first transplantation, but the frequency returned thereafter, leading to a second transplantation. As with the first transplantation, the subject experienced improvement in swallowing and blockage for some time after the second transplantation. However, since the frequency of EBD returned to the pre-transplantation level within 2 months, surgical resection of the stenotic esophagus was performed 4 months after the second transplantation. The effect of cell sheet transplantation was limited for this subject. During surgery, mobilization of the stenotic portion was difficult because of the scarring. However, the stenotic portion and the lower esophagus was removed, then esophageal reconstruction was performed using a total gastric tube. The post operative course was good. Although 2 EBDs were required for postoperative anastomotic stenosis, the stenotic symptoms disappeared thereafter for 3 years. Fortunately, the surgical treatment went well despite the high risk caused by the history of open thoracotomies, and a suture failure. However, it is desirable to be able to choose a treatment other than resection and anastomosis. In addition, in the resected esophageal stricture, fibrosis of the submucosa was noticeable and thickened. The thickness of the submucosal layer was 1.8 mm to 2.0 mm, which is 0.17 mm to 0.24 mm in the normal esophagus, suggesting that the repeated EBDs and healing could have been accumulation of inflammatory stimuli leading to severe refractory stenosis with thickening of the esophageal wall.

Subject 2 required EBDs every 3 months before transplantation, but after transplantation, he consumed a normal diet without EBD for more than 18 months, showing that the cell sheet transplantation treatment was effective. Even though change in the size of the lumen of the stenosis was not noticeable in contrast esophagography and endoscopic images, the subject experienced clear improvement in passage of food and drink after transplantation and was able to consume normal meals, suggesting that the stenotic site acquired some flexibility and expand during food passage. Based on these results, measuring the flexibility of the stenotic site may become a new index for evaluating treatment efficacy in the future.

Subject 3 had undergone EBD 2 or 3 times a year before transplantation, with adjustments in diet and duration of meals. However, after transplantation, he did not receive dilation for at least 9 months. In addition, the subject experienced a reduction in difficulty swallowing and blockage of food and drink, and the actual time required to eat was reduced by one-fourth, from 2 hours to 30 minutes. This subject also showed no significant change in the size of the lumen of the stenosis before and after implantation in imaging examinations, but since there was an improvement in both objective and subjective symptoms, it is assumed that the esophageal tissue may have gained flexibility, as in subject 2. However, since this subject is still in the follow-up period; therefore, the effect of the transplant treatment should be evaluated and confirmed later.

### Safety of cell sheet transplantation for post-anastomotic stenosis of CEA and CES

Since none of the 3 subjects showed any problems in physical or laboratory examinations, and because the epithelial sheets had passed quality control, sheet transplantation therapy for CEA and CES appears to be a safe and apparently less risky treatment compared to surgical removal of the stenotic esophagus.

### Differences in the effects of cell sheet transplantation between ECSS, CEA and CES

Cell sheet transplantation in the three CEA and CES patients in this study did not show the same dramatic effect as the transplants in ESCC patients. One reason for this could be the difference in the degree of fibrotic scarring at the transplant site. In the treatment of adult ESCC with ESD, which was the subject of the previous study by Okano et al, the esophageal mucosa alone or with the submucosa were dissected. On the other hand, in the initial surgical treatment of CEA and CES, which are the subjects of the current study, all layers of the esophageal wall were cut and sutured. Then, the EBDs were performed repeatedly for the post-anastomotic stricture to dilate the whole layer of the stenotic esophagus. Therefore, the depth of pre-existing damage in the esophageal wall at transplantation is far deeper and more severe in CEA and CES patients than in ECSS patients. In fact, histopathology of the resected esophageal stenosis of subject 1 revealed significant spreading of fibrosis and thickening of the submucosa. Significant fibrosis and thickening of the submucosal layer in resected refractory esophageal anastomotic strictures have been reported in other clinical studies as well [9]. This apparent difference in depth of injury or the degree of scarring of esophageal tissue appears to influence on the efficacy of cell sheet therapy when comparing the prevention of stenosis after ESD in adult ESCC with the prevention of restenosis after EBD of postoperative anastomotic stenosis in CEA and CES. The second reason for the different effect between CEA and CES vs ECSS is the difference in the size of the lumen at the transplantation site, which influences the technique used and the post-transplantation observations. ESCC patients are adults and have large esophageal lumens, which allows for a large working space for transplantation. On the other hand, CEA and CES patients are children and generally have smaller esophagi; in addition, the stenotic sites tend to collapse even right after EBD, causing technical difficulties in performing the transplantation accurately. For the best adaptation of the transplant sheets, they should be placed accurately to the targeted location, avoiding overlapping. Limited available space may therefore be a factor in the inability of the cell sheets to be fully effective in children. As a countermeasure, it is necessary to develop a new device that can accurately aid transplantation in very confined spaces, as well as new techniques or devices to keep the lumen dilated during surgery.

### Difference of the effect of cell sheet transplantation between subjects

The epithelial cell sheets used in this study were manufactured according to standard operating procedures (SOPs) and their quality was assured by quality control testing. Despite the equal quality of the epithelial cell sheets, the progress after transplantation differed between subjects. We believe that this is due to differences in the conditions of the esophageal stenosis between subjects. One possible difference is the degree of normal continuation of layers remaining in the stenotic tissue at the anastomosis: since the tension is generally very strong at the initial anastomosis site in CEA and CES, all or some of the layers of the esophagus may be partially separated postoperatively, instead of the full circumferential attachment of each layer with the lack of continuity of smooth muscle layer, as seen in subject 1. If the lack of layers is severe, it is assumed that the esophageal tissue will not heal normally after surgery, resulting in stenosis refractory to EBDs. In fact, it has been reported that cases of refractory anastomotic stenosis are more common in long-gap cases, in which the upper and lower esophagus are widely separated [1]. Second, repeated dilatation and restenosis may lead to severe scarring of the remaining submucosa and muscularis propria. In subject 1, accumulation of α-SMA positive myofibroblast indicates active scar forming regenerative reactions. The cumulative number of EBDs was another difference between subjects. It can be inferred that subject 1 had the most advanced scarring of the stenosis after more than a hundred EBDs. The third difference is the length of the stenosis. As seen in esophagography, the stenosis in subject 1 was considerably longer than in the other subjects. Since EBD causes lacerations equal in length to the stenosis, the longer the stenosis, the longer the extent of the laceration, resulting in the greater the extent of scarring. Fourth, the ratio of mucosal dissection caused by EBD to the circumferential diameter of the esophageal lumen also differed among subjects. Subjects 1 and 3 had full circumferential mucosal detachment at one region of the stenotic esophagus, whereas subject 2 only had detachment of the mucosa around the longitudinal laceration. This difference of the extent of mucosal detachment may be related to the length of the stricture and the degree of scarring. From these observations it can be inferred that the scarring of the esophageal stenosis tissue was less advanced in subjects 2 and 3 than in subject 1. The effectiveness of cell sheet transplantation was also higher in Subjects 2 and 3 than in Subject 1, suggesting that cell sheet transplantation may be more effective when the number of EBD procedures for postoperative anastomotic restenosis is low.

### Possible anti-fibrotic effect of cell sheet transplantation

Although the mechanism is not clear, transplanted epithelial cell sheets may act in some way to inhibit scarring of esophageal tissue or reduce scar tissue during the healing process of injury caused by EBD. As a result, the extent of pre-existing fibrosis or inflammation in the stenotic site may have influenced the clinical differences in efficacy after cell sheet transplantation between subjects observed in this study.

## Conclusions

Although progress after cell sheet transplantation differed among the three subjects, they all experienced better food passage for a longer time after the transplantation than they did after EBD without transplantation. The second subject showed clear effectiveness, with no requirement for EBD for an extended period of time after transplantation, confirming that cell sheet transplantation therapy is effective in some cases. This is a significant finding. In the future, it will be necessary to study more cases; develop technologies such as indices to evaluate the effectiveness of cell sheet transplantation therapy objectively and new devices to realize more accurate transplantation; identify cases in which the current therapy is effective; find the optimal timing for transplantation; and clarify the mechanism by which the current therapy improves stenosis.

## Data Availability

The datasets used and/or analyzed during the current study are available from the corresponding author on reasonable request.

## List of abbreviations

CEA: congenital esophageal atresia
CES: congenital esophageal stenosis
EBD: Endoscopic balloon dilatation
ESD: Endoscopic submucosal dissection
ESCC: Esophageal squamous cell carcinoma
CPF: Cell processing facility
SOP: Standard operating procedures
EDTA: Ethylenediamine tetraacetic acid tetrasodium
KCM: Keratinocyte culture medium

## Declarations

### Ethics approval and consent to participate

The entire study including the clinical intervention component and experiments with human cells and tissues was approved by the Institutional Review Board and Certified Special Committee for Regenerative Medicine at the National Center for Child Health and Development. Human cells in this study were utilized in full compliance with the Ethical Guidelines for Medical and Health Research Involving Human Subjects (Ministry of Health, Labor, and Welfare (MHLW), Japan; Ministry of Education, Culture, Sports, Science and Technology (MEXT), Japan). Informed consent to participate in this study was obtained from the parents of the patients.

### Consent for publication

Informed consent for publication was obtained from the parents of the patients.

### Authors’ contributions

AF, YF, and NK designed experiments. AF, YB, and NI collected tissue from the patient. RT fabricated cell sheets and performed quality control tests. AF, YF, and KA performed EBDs. YF, MA, and MM performed transplantation of cell sheets. AF, TM, MK, YY, MO, and KA performed follow-up examinations. TY performed histopathological examinations. MM contributed reagents, materials, and transplantation tools. AF, YF, and AU discussed the data and manuscript. AF, YF, and RT wrote this manuscript. All authors read and approved the final manuscript.

## Acknowledgements

We would like to express our sincere thanks to K. Ishikawa for providing expert technical assistance, to C. Ketcham for English editing and proofreading, to E. Suzuki for English writing, and to K. Saito for secretarial work.

## Supplemental Information

**Supplemental Figure 1. Fabrication and quality control tests for cultured autologous oral mucosal epithelial cell sheets**

Morphology of an autologous oral mucosa-derived epithelial cell sheet (left panels), morphology of oral mucosa cells before transport to the hospital where transplantation was performed (middle panels), and histogram of the percentage of epithelial cells in the cell sheet measured by flow cytometry detection of cytokeratin positive cells (right panels).

**Supplemental Table 1. Inspection items for epithelial cell sheet transplantation and their timing**

