## Supplementary figures and images for "Evaluation of safety and efficacy of autologous oral mucosa-derived epithelial cell sheet transplantation for prevention of anastomotic restenosis in congenital esophageal atresia and congenital esophageal stenosis: three case studies"

### Supplemental Figure 1

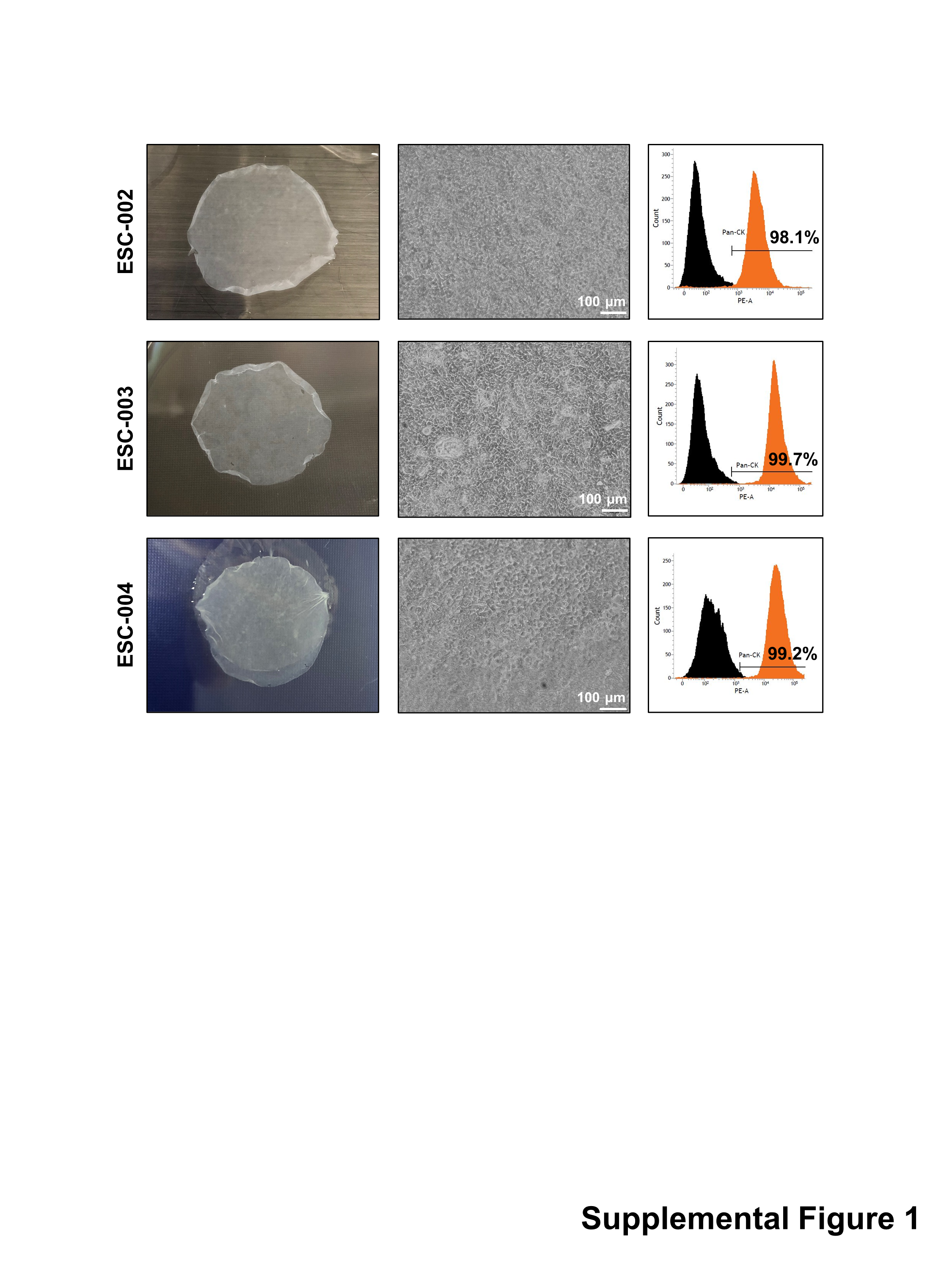
