## Supplemental Table 1 for "Evaluation of safety and efficacy of autologous oral mucosa-derived epithelial cell sheet transplantation for prevention of anastomotic restenosis in congenital esophageal atresia and congenital esophageal stenosis: three case studies"

| Observation/evaluation date | | Before treatment | Cell collection | Operation date | Day 1 after operation | Week 1 | Week 3 | Week 4 | Week 12 | Week 24 | Week 36 | Week 48 | Termination |
|---|---|---|---|---|---|---|---|---|---|---|---|---|---|
| Acceptable advance/delay in schedule | | 4 weeks in advance of operation date | 2 weeks in advance of operation date | 0 days | 1 day | ±2 days | ±1 week | ±1 week | ±4 weeks | ±4 weeks | ±4 weeks | ±4 weeks | - |
| Informed consent | | ○, ○ (Collection) | | ○ (Transplantation) | | | | | | | | | |
| Collection of blood and oral mucosa | | | ○ | | | | | | | | | | |
| Clinical condition (general) | Vital signs | ○ | | ○ | ○ | ○ | ○ | ○ | ○ | ○ | ○ | ○ | ○ |
| Clinical condition (stenosis) | Difficulty in swallowing, food stuck | ○ | | ○ | ○ | ○ | ○ | ○ | ○ | ○ | ○ | ○ | ○ |
| Clinical examination | Blood test | ○ | | | | ○ | (○) | ○ | ○ | | | ○ | ○ |
| | X-ray | ○ | | | | ○ | | ○ | ○ | | | ○ | ○ |
| Endoscopic examination | Presence of stenosis | ○ | | ○ | | ○ | | ○ | ○ | | | ○ | ○ |
| | Sheet fixing, epithelialization | | | ○ | | ○ | | ○ | ○ | | | ○ | ○ |
| Esophagography | Presence of stenosis | ○ | | | | | | ○ | ○ | | | ○ | ○ |
| Safety | Adverse event | ↔ | | | | | | | | | | | |

Supplemental Table 1. Inspection items for epithelial cell sheet transplantation and their timing
